# Study on the expression levels of antibodies against SARS-CoV-2 at different period of disease and its related factors in 192 cases of COVID-19 patients

**DOI:** 10.1101/2020.05.22.20102525

**Authors:** Jingyi Ou, Mingkai Tan, Haolan He, Haiyan Tan, Jiewen Mai, Yaoxiang Long, Xiaowen Jiang, Qing He, Ying Huang, Yan Li, Renshen Chen, Liya Li, Yaling Shi, Fang Li

**Affiliations:** Guangzhou Eighth People’s Hospital, Guangzhou Medical University, Guangzhou, China.; Department of Laboratory Medicine, Guangzhou Baiyun District Maternal and Child Health Hospital, Guangzhou, China.

## Abstract

**Background:** In 2020, the current outbreak of Coronavirus Disease 2019(COVID-19) has constituted a global pandemic. But the question about the immune mechanism of patients with COVID-19 is unclear and cause particular concern to the world. Here, we launched a follow-up analysis of antibodies against SARS-CoV-2 of 192 COVID-19 patients, aiming to depict a kinetics profile of antibodies against SARS-CoV-2 and explore the related factors of antibodies expression against SARS-CoV-2 in COVID-19 patient.

**Methods:** A total of 192 COVID-19 patients enrolled in the designated hospital of Guangzhou, Guangzhou Eighth People’s Hospital, from January to February 2020 were selected as the study cohort. A cohort of 130 COVID-19 suspects who had been excluded from SARS-CoV-2 infected by negative RT-PCR result and 209 healthy people were enrolled in this study. Detection of IgM and IgG against SARS-CoV-2 were performed by Chemiluminescence immunoassay in different groups.

**Results:** It has been found that the seroconversion time of IgM against SARS-CoV-2 in most patients was 5-10 days after the symptoms onset, and then rose rapidly, reaching a peak around 2 to 3 weeks, and the median peak concentration was 2.705 AU / mL. The peak of IgM maintained within one week, and then enters the descending channel. IgG seroconverted later than or synchronously with IgM, reaching peaks around 3 to 4 weeks.The median peak concentration was 33.998AU / ml,which was higher than that of IgM. IgM titers begins to gradually decrease after reaching the peak in the 4^th^ week, after the 8^th^ week, a majority of IgM in patient’s serum started to turn negative. On the contrary, titers of IgG began to decline slightly after the fifth week, and more than 90% of results of patients were positive after 8 weeks. Additionally, the concentration of antibodies positively correlated with the severity of the disease and the duration of virus exist in host.

**Conclusion:** We depict a kinetics profile of antibodies against SARS-CoV-2 in COVID-19 patients and found out that the levels of antibodies were related to the disease severity, age, gender and virus clearance or continuous proliferation of COVID-19 patients.

---

In December 2019, cases of unexplained pneumonia outbreak in Wuhan, China. Later, it was found that the pathogen causing Coronavirus Disease 2019(COVID-19) was a novel coronavirus (SARS-CoV-2). The current outbreak of COVID-19 has constituted a global pandemic.As of May 18,2020,a total of 4.8 million confirmed cases and more than 310000 death cases have been reported over the world. The clinical manifestation of SARS-CoV-2 infected people have varied from no symptom to severe pneumonia. Furthermore, asymptomatic cases could be the virus carrier and spread to more health people. Meanwhile, the severe patients may have acute respiratory distress syndrome, respiratory failure and required monitoring and treatment in Intensive Care Unit (ICU).Therefore the epidemic spread of SARS-CoV-2 threats to the public health and brings a negative impact on the social economy.

Along with the obtainment of complete SARS-CoV-2 genome sequences from COVID-19 patients in Wuhan [1], real-time reverse transcription PCR(RT-PCR) assay started to be used in most laboratories as a “golden standard” for COVID-19 diagnosis. Nevertheless, the positive rate of RT-PCR for SARS-CoV-2 was only about 50% [2], and RT-PCR assay may need high standard examiner and sophisticated equipment.

Some researchers have found out that the titer of SARS-CoV-2 antibodies got dynamic increased in the serum of patients with COVID-19 [3-5], different kinds of serologic testing kit have been developed. Chemiluminescence immunoassay has the advantages of higher sensitivity and specificity, more stable dectection system, and higher laboratory biosafety than other immunoassays. Here, we launched follow-up analysis of antibodies against SARS-CoV-2 of 129 COVID-19 patients, and among them 94 rehabilitation patients has been recruited for testing the titers of antibodies, aiming to analyze the expression levels of antibodies against SARS-CoV-2 at different period of disease and explore its related factors in COVID-19 patient.

## Methods

### Data Collection

A total of 192 COVID-19 patients enrolled in the designated hospital of Guangzhou, Guangzhou Eighth People’s Hospital, from January to February 2020 were selected as the study cohort. The confirmed diagnoses of COVID-19 patients were confirmed by RT-PCR assay. The degree of infection severity may vary from mild to severe. Based on the Guidelines for Diagnosis and Treatment of Novel Coronavirus Pneumonia (Seventh version), released by the National Health Commission of China, the severe patient should follow at least one of the criteria:(1)shortness of breath(respiratory rate ≥ 30 breaths per minute);(2)arterial oxygen saturation(resting status) ≤ 93%; or (3) the ratio of partial pressure of oxygen to fraction of inspired oxygen(PaO_2_/FiO_2_) ≤ 300mmHg. Critical patients should meet with at least one of the followings: (1)Respiratory failure occurred and mechanical ventilation was required; (2)Shock occurred;(3) Multiple organ failure occurred, patients were required monitoring and treatment in ICU. To evaluate the differential diagnosis ability of the chemiluminescence kit,a cohort of 130 COVID-19 suspects who had been excluded from SARS-CoV-2 infected by negative RT-PCR result were enrolled in this study[Median age (IQR):24(21-32)years;Male:59.2%]. A cohort of 209 healthy people who performed the physical examination in Guangzhou Baiyun District Maternal and Child Health Hospital (excluded from COVID-19, respiratory system infections, cardiovascular system disease, hepatitis, immune system disease)were selected as the reference sample group and negative control group. The median age of the healthy people was 49(IQR, 32-56years) years and 47.8% was male. All patients in this study have signed the informed consent, and the experimental protocol was approved by the Ethics Committee of Guangzhou Eighth People’s Hospital(No.20200547).

### Detection of IgM and IgG against SARS-CoV-2 by Chemiluminescence immunoassay

Approximately 3-5 milliliter of blood samples were collected from examinees in different groups, then then they were centrifuged at 1509.3×g for10 minutes to separate serum. The collected samples were tested on the **MAGLUMI**^®^ 800 (Shenzhen New Industries Biomedical Engineering Co.,Ltd [Snibe], China) chemiluminescent analytical system(CLIA),according to the manufacturer’s instructions.The CLIA for IgM and IgG detection were based on the chemiluminescent immunocapture method and immunoindirect method respectively.The recombinant antigens contains the nucleocapsid protein and spike protein of SARS-CoV-2. N-(4-Aminobutyl)-N-ethylisoluminol was used as a chemiluminescent marker in these methods. According to the manufacturer’s instruction, the SARS-CoV-2 IgM and IgG cut-off are both 1.0 AU/mL.

### Statistical analysis

Continuous variable data were demonstrated by mean (standard deviation) or median (interquartile range), while categorical variables data were expressed by frequency and percentage. Parametric test (*t* test) and nonparametric test (Mann-Whitney *U* test) were used for continuous variables with or without normal distribution, respectively. Chi-square test(*χ*^2^ test) was used for categorical variables. Multivariate analysis of antibodies level was using multiple linear regression equations. The relation between levels of antibodies and ages was analyzed by Pearson’s correlation*. P* value of less than 0.05 is regarded as statistical significance. Statistical analysis were performed by SPSS version 22.0(IBM SPSS, Chicago, Illinois) and GraphPad Prism version 5.0(Graphpad Software, Inc., CA, USA).

### Data availability

The data that support the findings of this study are available from the corresponding author upon reasonable request.

## Results

### Demographic and clinical characteristics of COVID-19 patients

Total of 192 COVID-19 patients with clear clinical information were recruited in the study group. The median age of the study group was 52 (IQR, 36-62 years) years, and among of them 45.8% were male. 156 patients with mild or common symptom were assigned to the non-severe group, while 36 patients who deteriorated to severe or critical state were enrolled into the severe group. The baseline characteristics of COVID-19 patients in two groups are demonstrated in Table1. The median age of severe group was older than that of non-severe group(*P*<0.001), and the severe group has longer hospitalization (*P*<0.001). Male patients were more likely to deteriorated to severe or critical COVID-19 than female patients. 48.6% of patients with basic disease were in severe group while 36.9% were in non-severe group. Nevertheless, there were no statistical differences in basic disease and Body Mass Index (BMI) between the two groups. As of the end of the writing of the article, 4 patients have not been discharged because of continuously treatment of basic disease. The hospitalization time of them were both more than 110 days.

**Table 1.** Baseline Characteristics of patients with Coronavirus Disease 2019.

|  | **Non-severe**<br>COVID-19(n=156) | **Severe**<br>COVID-19(n=36) | **P** |
| --- | --- | --- | --- |
| **Age, y, X±SD** | 49±15 | 60±13 | <0.001 |
| **Sex** |  |  |  |
| Male | 65(41.7%) | 23(63.9%) | 0.016 |
| Female | 91(58.3%) | 13(36.1%) |  |
| **Interval between symptom onset to admission (days)** | 5±3 | 5±3 | 0.471 |
| **Basic disease, No.(%)** |  |  |  |
| No | 98(63.5%) | 19(51.4%) | 0.266 |
| Yes | 58(36.9%) | 17(48.6%) |  |
| Hypertension, No.(%) | 27(17.3%) | 11(30.6%) | 0.072 |
| Diabetes, No.(%) | 9(5.8%) | 3(8.3%) | 0.567 |
| Hyperlipidemia, No.(%) | 3(1.9%) | 1(2.7%) | 0.746 |
| Hepatitis, No.(%) | 8(5.1%) | 2(5.6%) | 0.917 |
| **BMI, kg/m<sup>2</sup>** | 23.9±3.7 | 24.9±5.0 | 0.453 |

Data are presented as no.(%) unless otherwise indicated.

### Establishment of Relative Light Unit (RLU) cutoff value of the chemiluminescence kit for COVID-19 diagnosis

A cohort of 209 healthy people who performed the physical examination in Guangzhou Baiyun District Maternal and Child Health Hospital were selected as the reference sample group. Serological test was performed in the reference sample group using the chemiluminescence kit. Base on the Clinical and Laboratory Standards Institute(CLSI)EP28-A3c protocol[6],we used non-parametric method to calculate the 95% confidence interval of RLU in the reference population. As a result of the calculation, the RLU cutoff value of IgM and IgG against SARS-CoV-2 in our laboratory were 5000 and 9677 respectively, which were determined by the upper 95% confidence interval limit. This is very close to the luminescence threshold of IgM and IgG (5000 and 9000) declared by the manufacturer. The cutoff value belonged to IgM and IgG respectively both equivalented to 1.0 AU/mL.

### Test performance on chemiluminescence kit of antibodies against SARS-CoV-2

The serological tests of 192 COVID-19 patients, 130 COVID-19 suspects who had been excluded from COVID-19 infection, and 209 healthy people were performed using the chemiluminescence kit. The positive test rate of IgM (81.8%) were significantly lower than that of IgG (93.2%) in COVID-19 confirmed case(*P*<0.001).The false positive cases of IgM and IgG in negative control group were both at a lower rate. The chemiluminescence kit shown an excellent anti-interference ability in diagnosing COVID-19 for the extremely low false positive rate of COVID-19 suspect group. The detailed data was shown in Table 2.

**Table 2.** Test performance data on chemiluminescence kit of IgM/IgG antibodies against SARS-CoV-2.

|  | **IgM** | **IgG** | **P** |
| --- | --- | --- | --- |
| **COVID-19 confirmed group**<br>(n=192) |  |  |  |
| Positive | 157(81.8%) | 179(93.2%) | 0.001 |
| Negative | 35(18.2%) | 13(6.8%) | - |
| **COVID-19 suspect group**<br>(n=130) |  |  |  |
| Positive | 1(0.8%) | 1(0.8%) | 1.000 |
| Negative | 129(99.2%) | 129(99.2%) | - |
| **Negative control group**<br>(n=209) |  |  |  |
| Positive | 5(2.4%) | 4(1.9%) | 0.736 |
| Negative | 204(97.6%) | 205(98.1%) | - |

### Kinetics of IgM and IgG against SARS-CoV-2 in COVID-19 patients

To investigate the kinetics of IgM and IgG against SARS-CoV-2 in COVID-19 patients, a total of 1423 specimens were collected from 192 COVID-19 patients at different days after symptom onset during hospitalization and rehabilitation period. As the Tab.3 shown, the seroconversion time of IgM against SARS-CoV-2 in most patients was 5-10 days after the symptoms onset, and then rose rapidly, reaching a peak around 2 to 3 weeks, and the median peak concentration was 2.705 AU/mL. The peak of IgM maintained within one week, and then enters the descending channel. IgM has not been detected in one third of the patients about 5 weeks after the symptoms onset, and IgM has not been detected in more than half of the patients around 8 weeks. IgG seroconverted later than or synchronously with IgM, reaching peaks around 3 to 4 weeks. The median peak concentration was 33.998AU / ml, which was higher than that of IgM. The peak duration last for a long time, about 1-2 weeks, and then entered into a plateau or slowly decreasing, the duration can be as long as several weeks. IgG against SARS-CoV-2 could be detected in 96% of patients at about 8 weeks after the symptoms onset with high concentration.

Additionally, we also found patients with delayed seroconversion. For example, a 27 years old female patient(No.95,shown in Figure 4L) whose IgG was positive after being returned to the hospital for 33 days after discharge. It is now 56 days after the disease onset. This patient has no basic diseases. IgM and IgG of 13 patients were not detected, while 12 patients were observed during hospitalization to rehabilitation period, and only one patient remained negative during hospitalization, unfortunately missing observation in rehabilitation period. All of them are non-severe patients, with average 4 days from symptoms onset to admission, and 3 patients with asymptomatic infection (CT no pneumonia manifestation during hospitalization, all RT-PCR results were negative) and the other 10 patients were mild or common patients. The median age of them was 36 years old, and 5 of them were male. A Tpatient(No.69,shown in Figure 7J) had an upward trend of IgM concentration, but because of the longer detection interval, the IgM lost the seroconversion detection point. The patient (NO.78) who was shown in Figure 7K, has fewer monitoring times and longer time intervals between tests during the entire observation period, so it is not a complete observation case. Therefore, these patients with negative antibody tests do not necessarily produce no antibodies, but miss the best observation time.

**Figure 1.**
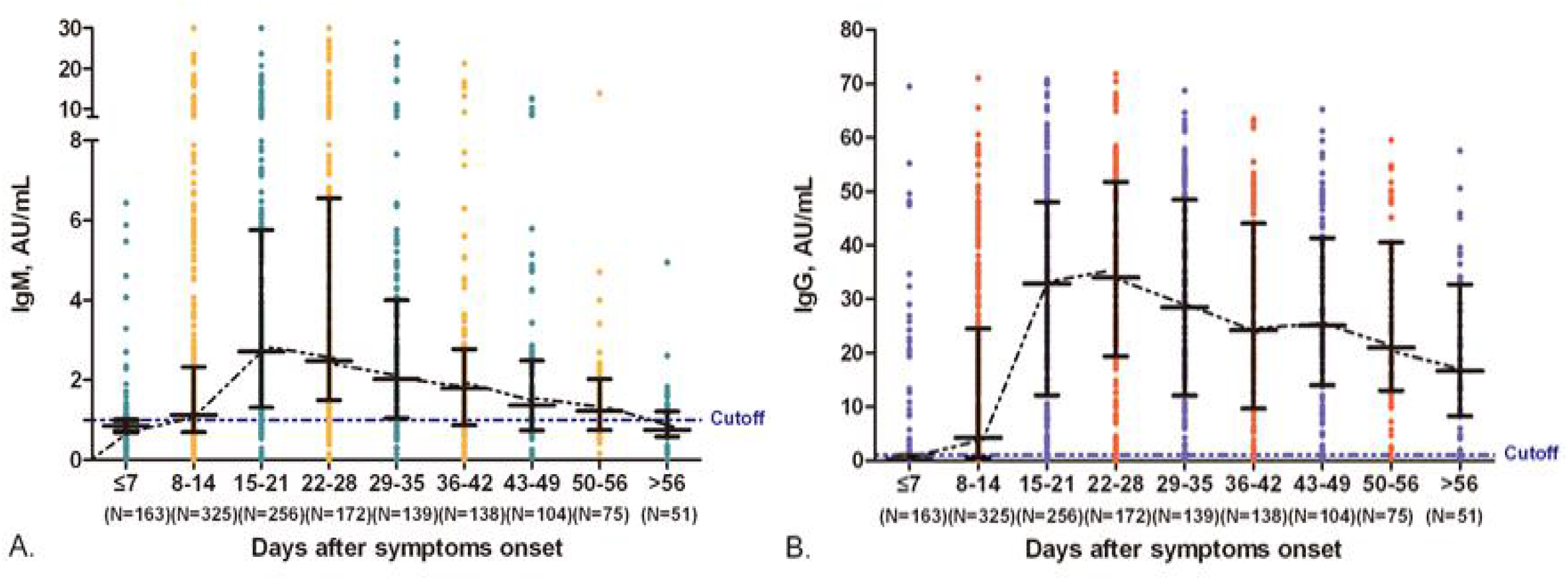
Kinetics of IgM and IgG against SARS-CoV-2 in COVID-19 patients.

**Figure 2.**
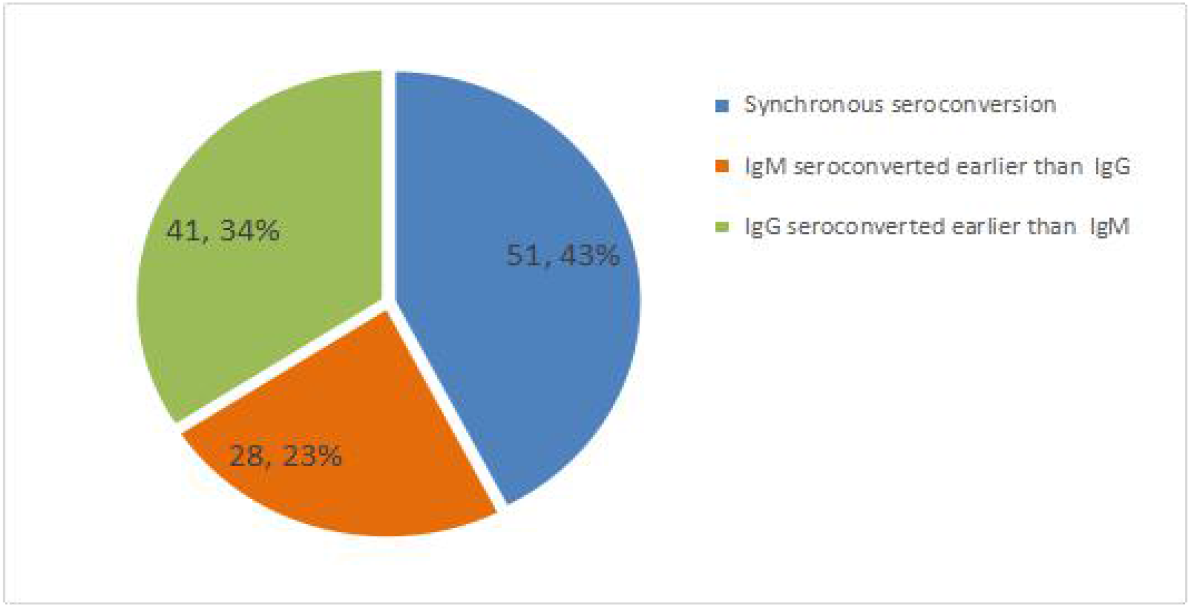
Pie chart of the seroconversion time of the IgM and IgG in COVID-19 patients.

**Figure 3.**
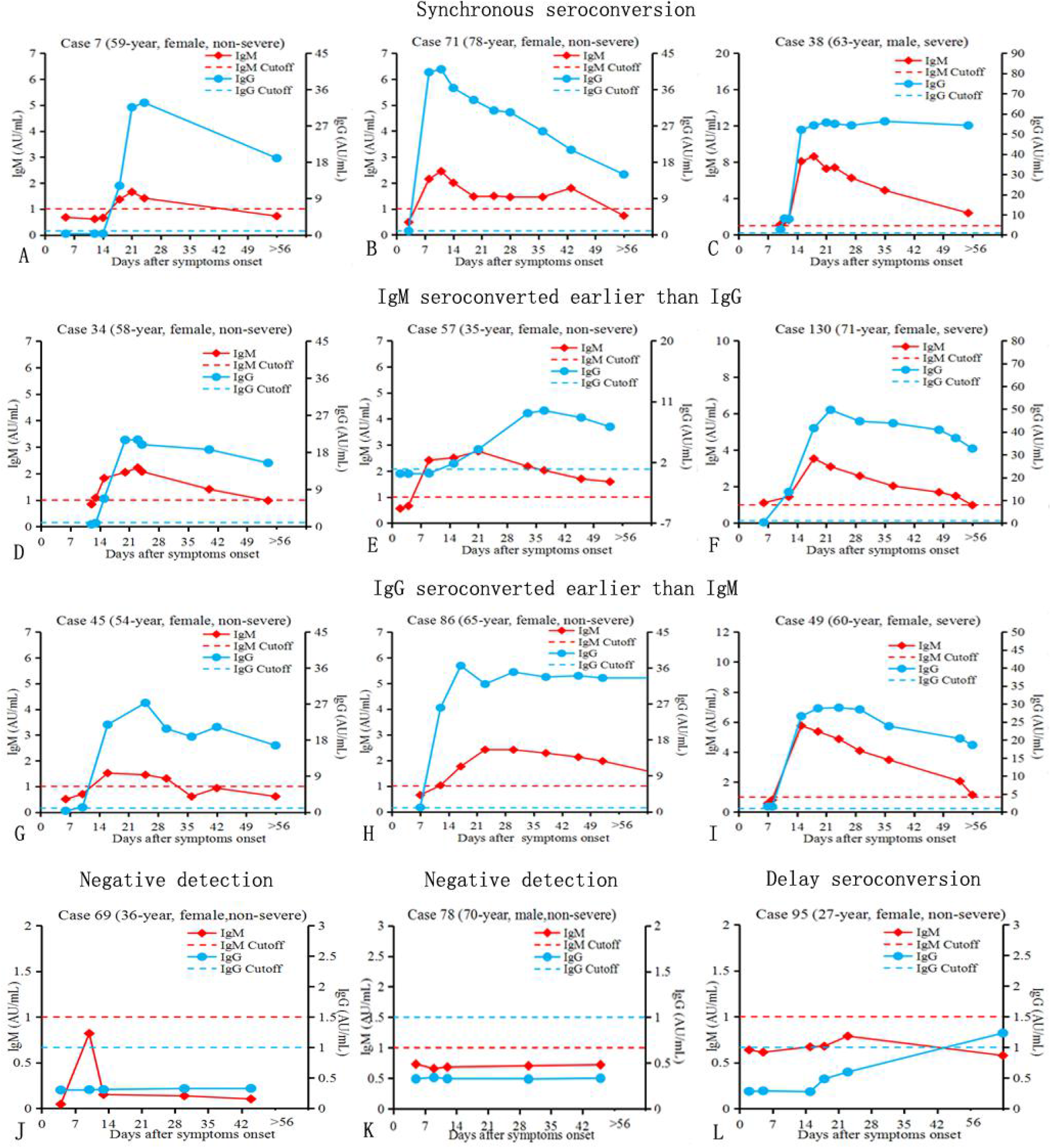
Typical example of the different seroconversion types.

**Figure 4.**
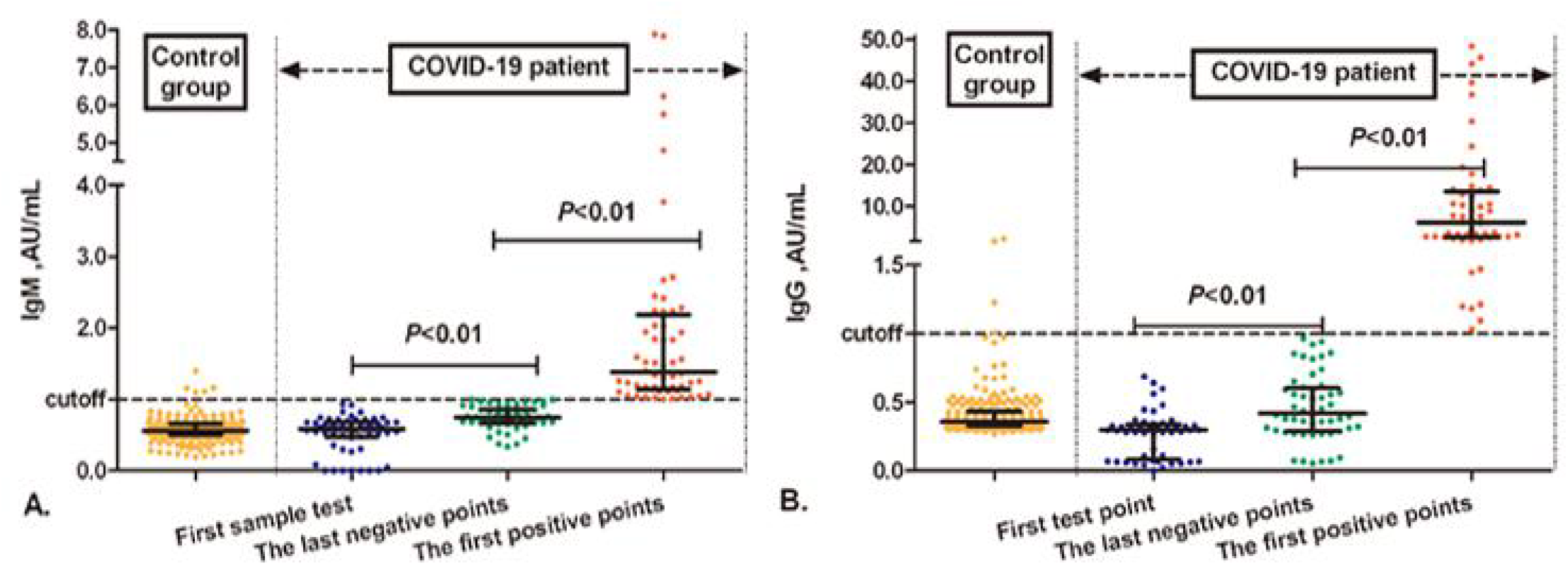
Difference in antibodies against SARS-CoV-2 expression between negative control group and COVID-19 confirmed group.

**Figure 5.**
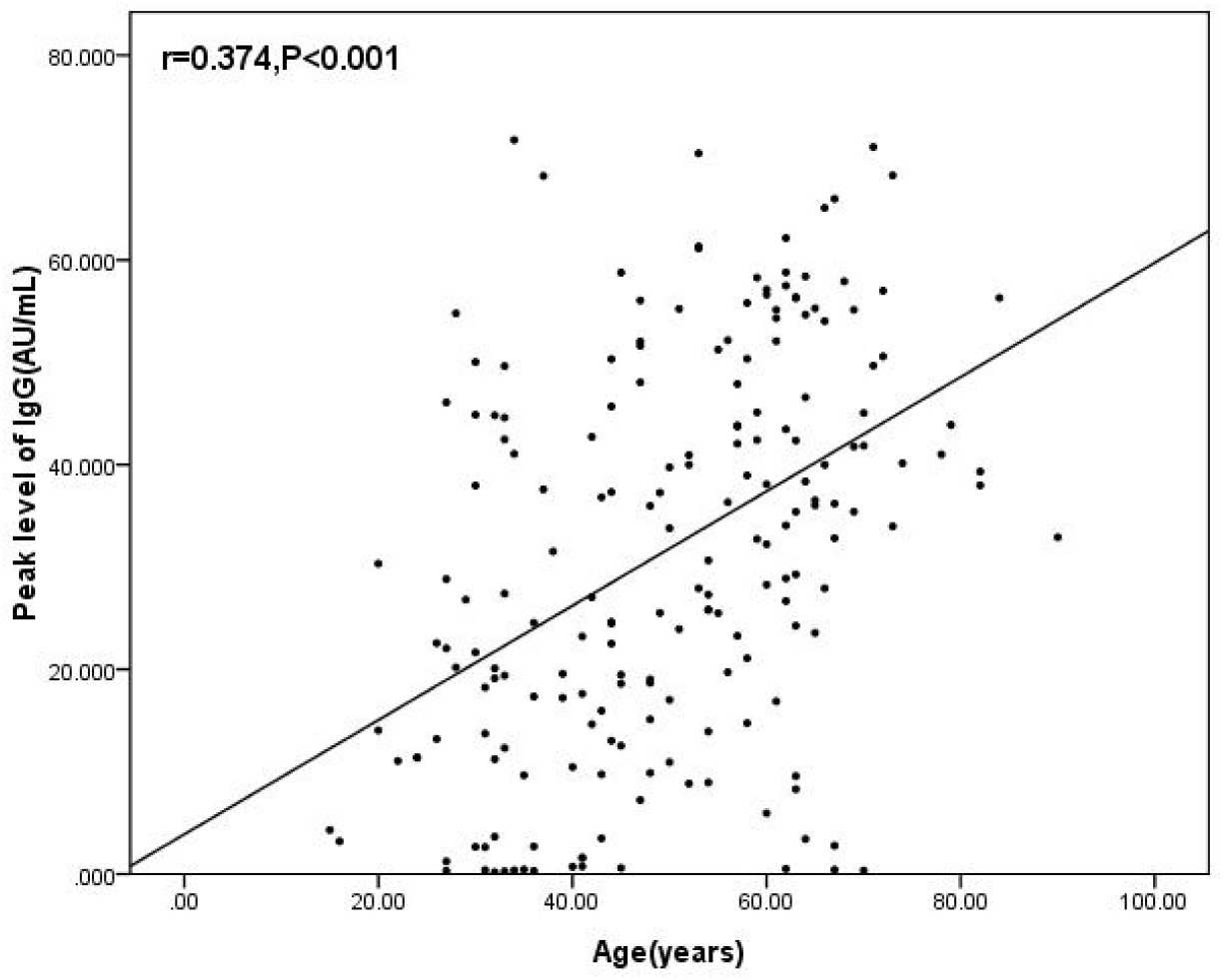
The Correlation between age and the concentration of IgG(AU/mL)

**Figure 6.**
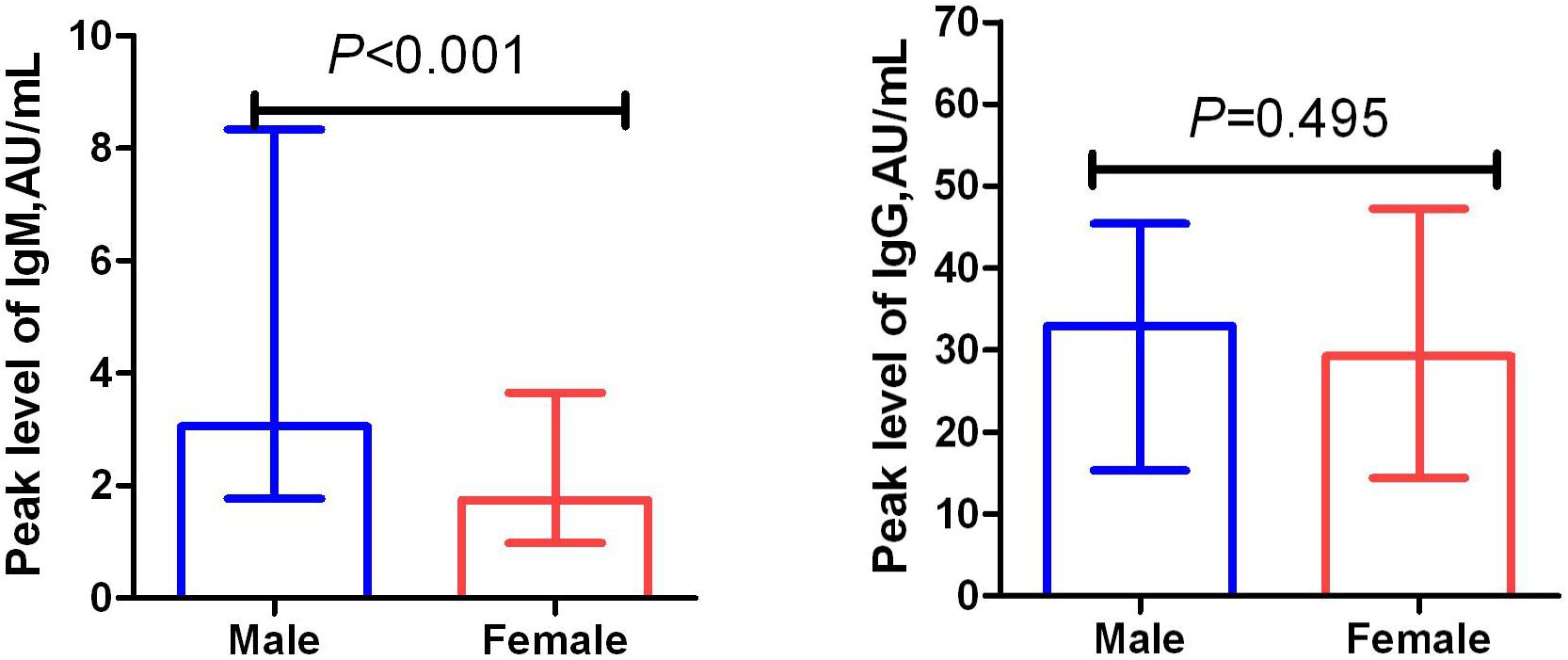
Expression levels of antibodies against SARS-CoV-2 response in different gender patients.

**Figure 7.**
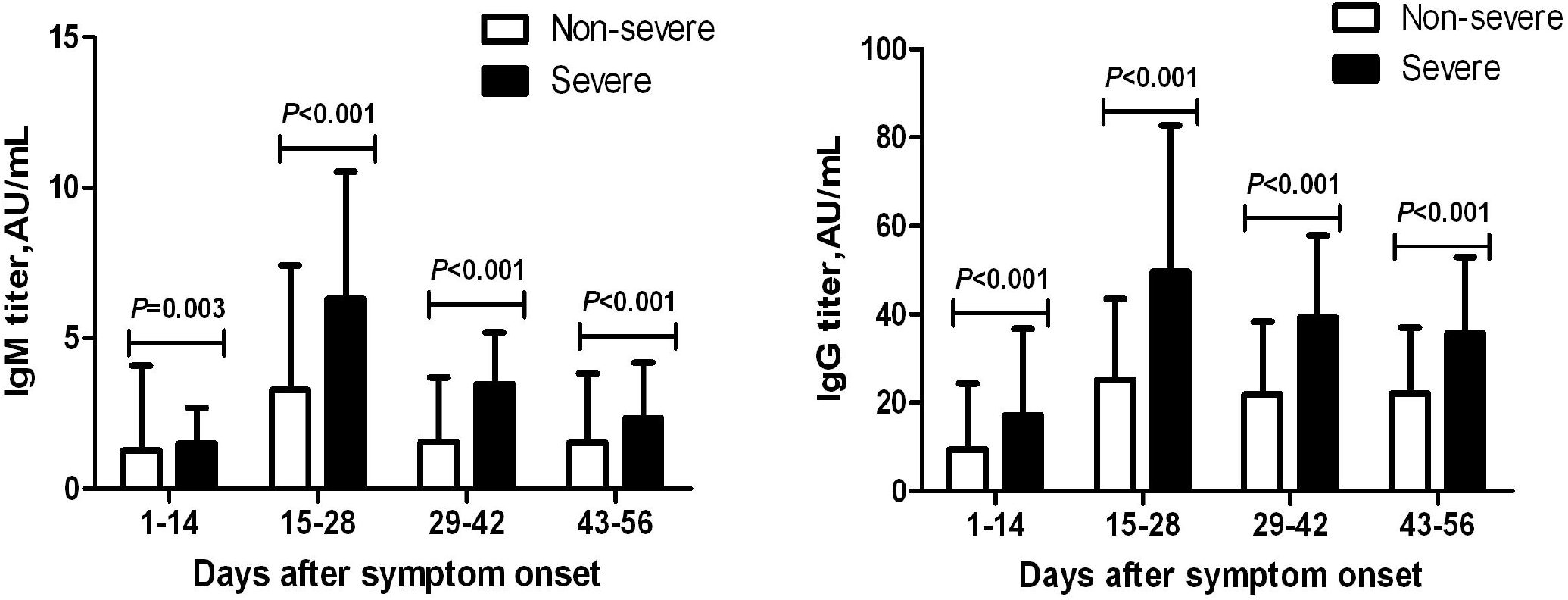
Kinetics of IgM and IgG against SARS-CoV-2 in non-severe and severe COVID-19 patients.

### Difference in antibodies against SARS-CoV-2 expression between negative control group and COVID-19 confirmed group

We sorted out the detection results of 3 time points (the first test point, the last negative point and the first positive point).The results of 3 time points were compared with the results of the control group, we found that the concentration of antibodies in patients before the seroconversion was higher than that of healthy population, with an increasing trend. It indicated that the antibodies of COVID-19 patients began to produce early in the infection, however the concentration of them have not reached the threshold we set so couldn’t be detected. Therefore, COVID-19 suspects whose concentration of antibodies are below “sea level” cannot be judged uninfected by only one test. The serological test of COVID-19 suspects should be dynamically observed multiple times to find out whether the antibody has an upward trend.

In order to analyze the difference between the concentration of antibodies before seroconversion in COVID-19 patients and healthy population, 56 cases of COVID-19 patients with at least two serological tests before IgM and IgG seroconversion were selected to the analysis. The serological tests were subdivided into 3 time points: the first test point (median days after onset 4 days), the last negative point (median days after onset 9 days)and the first positive points(median days after onset 13 days). The results showed that the concentration of IgM in the last negative test points was higher than that of control group. Moreover, IgM of COVID-19 patients started to increase in the early stage of disease, which was more obvious than that of IgG, but the concentration of IgG was significantly higher than that of IgM after seroconversion (Tab 4, Fig 4).

**Table 3.**
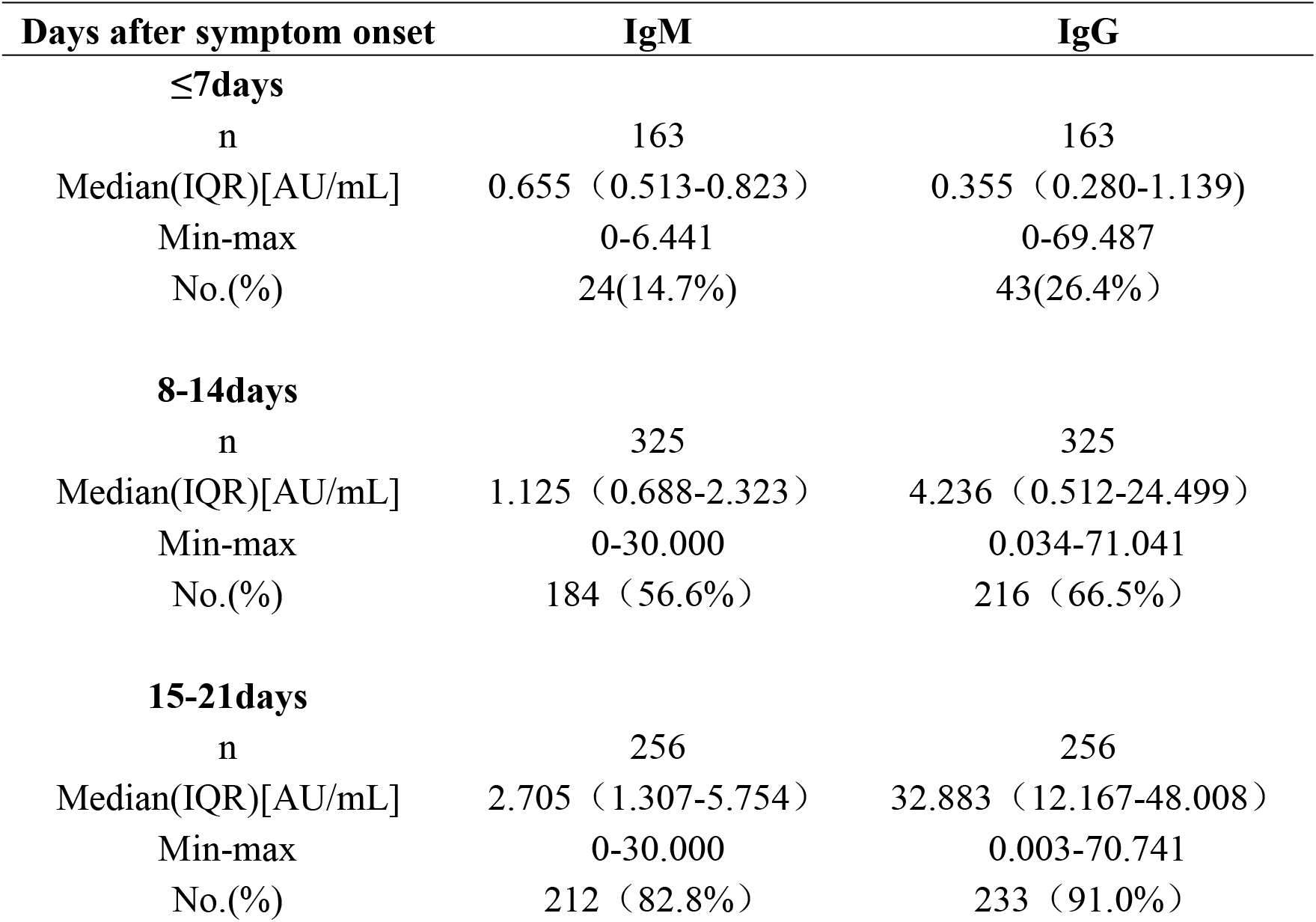
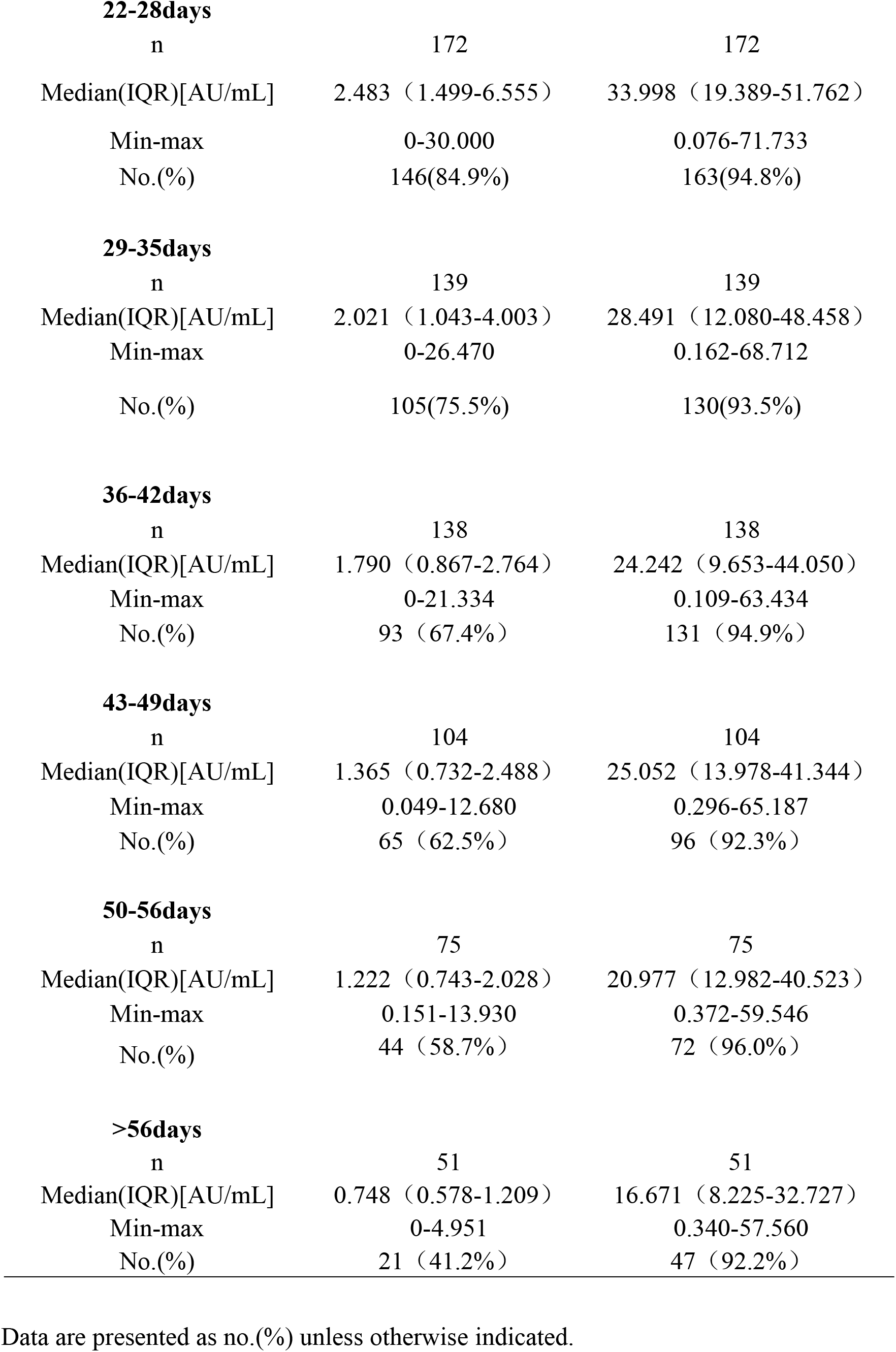
Descriptive statistics of IgM and IgG against SARS-CoV-2 of 192 COVID-19 patients(divided into 9 groups based on the days after symptom onset)

| **Days after symptom onset** | **IgM** | **IgG** |
| --- | --- | --- |
| **≤7days** |  |  |
| n | 163 | 163 |
| Median(IQR)[AU/mL] | 0.655 (0.513-0.823) | 0.355 (0.280-1.139) |
| Min-max | 0-6.441 | 0-69.487 |
| No.(%) | 24(14.7%) | 43(26.4%) |
| **8-14days** |  |  |
| n | 325 | 325 |
| Median(IQR)[AU/mL] | 1.125 (0.688-2.323) | 4.236 (0.512-24.499) |
| Min-max | 0-30.000 | 0.034-71.041 |
| No.(%) | 184 (56.6%) | 216 (66.5%) |
| **15-21days** |  |  |
| n | 256 | 256 |
| Median(IQR)[AU/mL] | 2.705 (1.307-5.754) | 32.883 (12.167-48.008) |
| Min-max | 0-30.000 | 0.003-70.741 |
| No.(%) | 212 (82.8%) | 233 (91.0%) |
| **22-28days** |  |  |
| n | 172 | 172 |
| Median(IQR)[AU/mL] | 2.483 (1.499-6.555) | 33.998 (19.389-51.762) |
| Min-max | 0-30.000 | 0.076-71.733 |
| No.(%) | 146(84.9%) | 163(94.8%) |
| **29-35days** |  |  |
| n | 139 | 139 |
| Median(IQR)[AU/mL] | 2.021 (1.043-4.003) | 28.491 (12.080-48.458) |
| Min-max | 0-26.470 | 0.162-68.712 |
| No.(%) | 105(75.5%) | 130(93.5%) |
| **36-42days** |  |  |
| n | 138 | 138 |
| Median(IQR)[AU/mL] | 1.790 (0.867-2.764) | 24.242 (9.653-44.050) |
| Min-max | 0-21.334 | 0.109-63.434 |
| No.(%) | 93 (67.4%) | 131 (94.9%) |
| **43-49days** |  |  |
| n | 104 | 104 |
| Median(IQR)[AU/mL] | 1.365 (0.732-2.488) | 25.052 (13.978-41.344) |
| Min-max | 0.049-12.680 | 0.296-65.187 |
| No.(%) | 65 (62.5%) | 96 (92.3%) |
| **50-56days** |  |  |
| n | 75 | 75 |
| Median(IQR)[AU/mL] | 1.222 (0.743-2.028) | 20.977 (12.982-40.523) |
| Min-max | 0.151-13.930 | 0.372-59.546 |
| No.(%) | 44 (58.7%) | 72 (96.0%) |
| **>56days** |  |  |
| n | 51 | 51 |
| Median(IQR)[AU/mL] | 0.748 (0.578-1.209) | 16.671 (8.225-32.727) |
| Min-max | 0-4.951 | 0.340-57.560 |
| No.(%) | 21 (41.2%) | 47 (92.2%) |

Data are presented as no.(%) unless otherwise indicated.

**Table 4.** The descriptive data of antibodies against SARS-CoV-2 in negative control group and COVID-19 patients.

|  | Negative control group<br>(n=209) | COVID-19 patients (n=56) |  |  |
| --- | --- | --- | --- | --- |
|  |  | First test points | The last negative test points | The first positive test points |
| Median days after symptoms onset | - | 4 | 9 | 13 |
| IgM,Median(IQR),AU/mL | 0.561(0.506-0.657) | 0.585(0.470-0.679) | 0.747(0.663-0.855)** | 1.383(1.138-2.183)** |
| IgG,Median(IQR),AU/mL | 0.356(0.328-0.434) | 0.297(0.082-0.337) | 0.416(0.290-0.604) | 6.027(2.543-13.541)** |

Note:Compared to the negative control group,\*\* $P<0.01$

### Related factors of antibodies against SARS-CoV-2 expression in COVID-19 patients

To screen out the related factors of antibodies against SARS-CoV-2 expression in COVID-19 patients, multiple linear regression equations were used to analysis the correlation. The selected variables are Age, gender, disease severity, basic disease and BMI were selected to be the variables. The results showed that the concentration of IgM in COVID-19 patients was related to gender and disease severity (*P* <0.01), and the concentration of IgG was related to age and disease severity (*P* <0.001)(Tab 5,6),. The result of univariate analysis of relevant factors, we found that IgG concentration showed a low level of correlation with age (r = 0.374, *P* <0.01)(Figure 5).The IgM concentration of male patients[(Median:3.054, IQR:1.771-8.335)AU/mL)] was higher than that of female patients[((Median:1.743, IQR:0.981-3.652)AU/mL](*P* <0.001) (Figure 6). Comparing the antibody levels of non-severe and severe patients at different stage of disease, it was found that no matter at what stage of the disease, the levels of IgG and IgM in the severe group were higher than those in the non-severe group. (Figure 7)

**Table 5.** Multivariate analysis of IgM level.

| Model | B | Std. Error | *t* | *P* |
| --- | --- | --- | --- | --- |
| Age | .004 | .031 | .138 | .891 |
| Gender | -2.635 | .907 | -2.906 | .004 |
| Disease severity | 3.328 | 1.205 | 2.761 | .006 |
| Basic disease | -.994 | .951 | -1.046 | .297 |
| BMI | .078 | .112 | .697 | .487 |

**Table 6.**
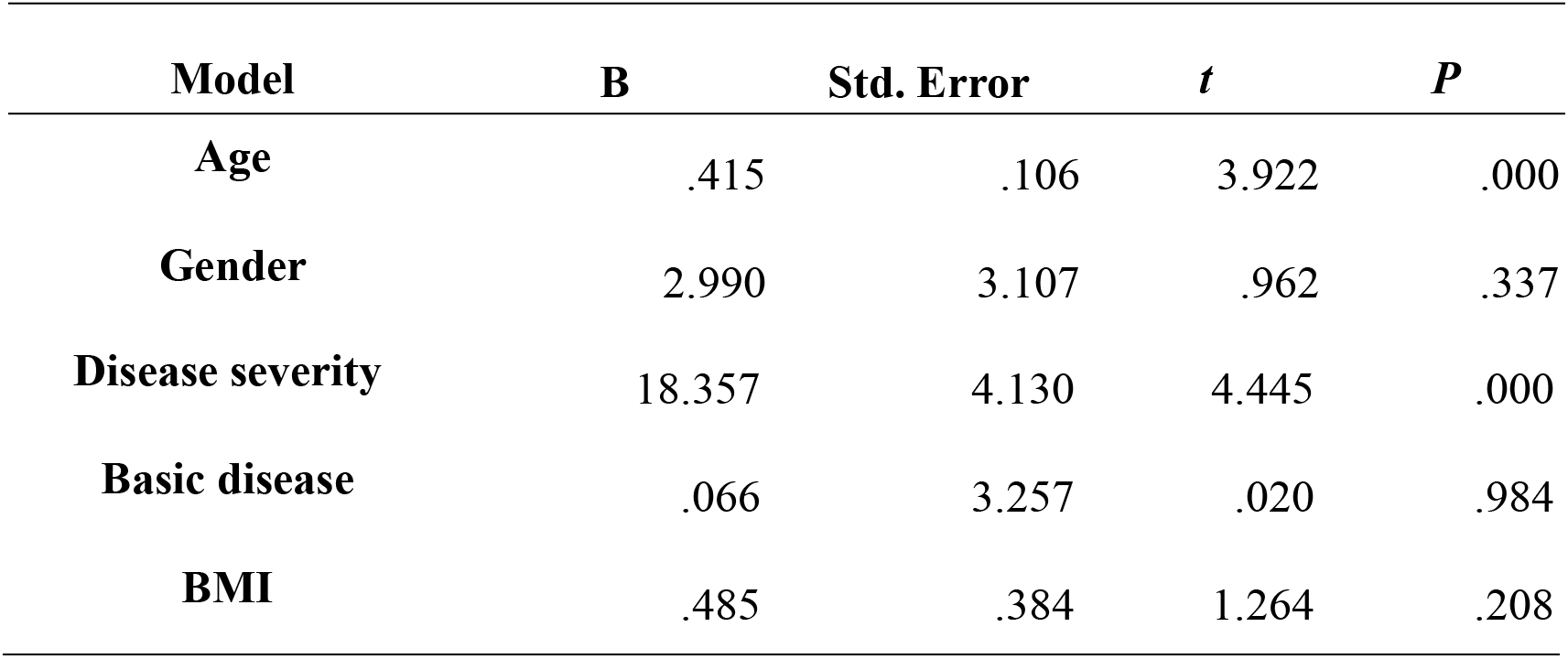
Multivariate analysis of IgG level.

| Model | B | Std. Error | *t* | *P* |
| --- | --- | --- | --- | --- |
| Age | .415 | .106 | 3.922 | .000 |
| Gender | 2.990 | 3.107 | .962 | .337 |
| **Disease severity** | 18.357 | 4.130 | 4.445 | .000 |
| **Basic disease** | .066 | 3.257 | .020 | .984 |
| **BMI** | .485 | .384 | 1.264 | .208 |

### The continuously results of RT-PCR and serological test in 4 cases of COVID-19 patients

We selected two patients with continuously serological test and RT-PCR results in the course of disease from non-severe group and severe group, aiming to explore the relationship between antibody and virus clearance or continuous proliferation in COVID-19 patients. The detailed information of each case was shown in Figure 8. We found that the IgG concentration of two non-severe patients reached a peak at 15-22 days after symptoms onset. The IgG concentration rose by more than 4 times, and then began to gradually decline. The result of RT-PCR at this time also began to turn negative and remained negative until discharge. The IgG concentration of two severe patients reached a peak (above 40 AU/mL) 15-17 days after the symptoms onset. During the subsequent observation period, the IgG concentration has remained at a high level, while the viral nucleic acid were continuously detected also in the observation period.

**Figure 8.**
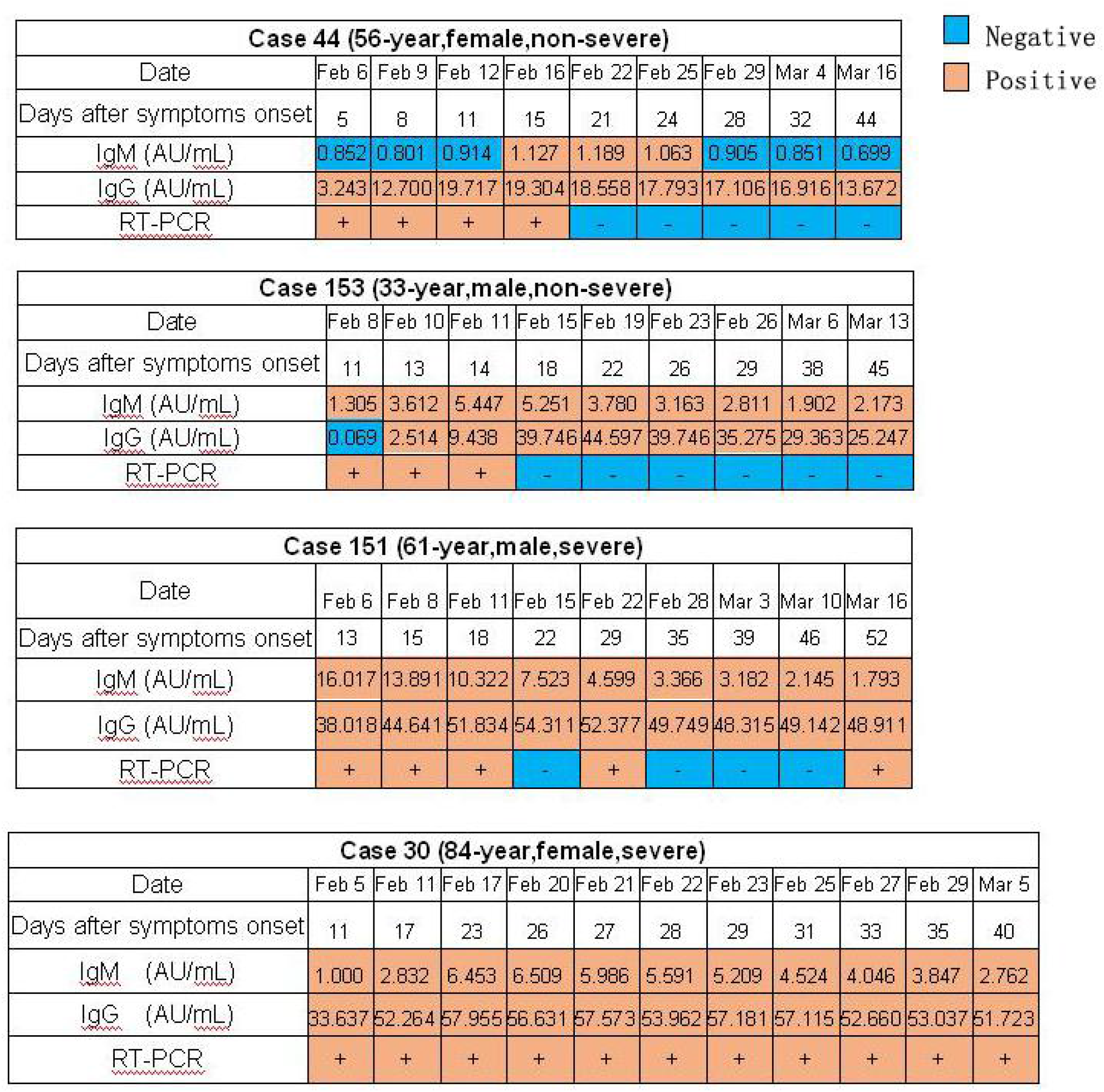
Four patients in the non-severe group and the severe group respectively with conducted follow-up analysis during the hospitalization.

## Discussion

According to the reports by World Health Organization, as of May 18, 2020, there have been more than 4.8 million confirmed cases of COVID-19 in the world. The question about the immune mechanism of patients with COVID-19 is of particular concern to the world. Hoping to understand the process of antibodies production in patients infected with SARS-CoV-2, and explore the related factors to the levels of antibody, total of 192 COVID-19 patients admitted to Guangzhou Eighth People’s Hospital were selected for serological follow-up analysis. Most of them started to be observed within two days of admission. The longest follow-up time was 81 days after rehabilitation and discharge, and the longest hospitalization time for discharged patients was 45 days. Two patients are still in hospital.

SARS-CoV-2 viral genome open reading frame (ORF) contains spike protein and nucleocapsid protein, which were considered to be the potential specific antigens for COVID-19 diagnostics in some studies [7, 8]. We used a chemiluminescence kit from Shenzhen New Industries Biomedical Engineering Co., Ltd [Snibe], China. In order to increase the detection rate of antibodies against SARS-CoV-2, the recombinant antigens contains spike protein and nucleocapsid proten simultaneously in this chemiluminescence kit.

Since the kit has to consider the problem of false positive rate in non-endemic areas when it is being developed, its declared positive judgment value may not be applicable to our regions and laboratories. The unsuitable positive judgment value will directly affect the sensitivity and specificity of the kit. Therefore, we decided to verify or reset the judgment value by a large sample. We randomly selected 209 healthy subjects as the reference control group, and used the same batch of reagents in our laboratory to perform serological testing on the control group samples. The upper limit of the 95% confidence interval of the luminescence value is compared with the positive interpretation value of the kit. Then the positive judgment value of the kit tested was determined in our laboratory. According to the positive judgment value, the kit has a higher detection rate of IgM and IgG against SARS-CoV-2, and the false positive rate is lower in healthy people (2.4% and 1.9%). At the same time, we conducted antibody monitoring on 130 suspected patients who were finally excluded, and only one patient had positive result, which means that combined antibody testing can have important value in the elimination of suspected patients.

In order to observe the production and changes of SARS-CoV-2 antibodies in patients with COVID-19, a statistical analysis of the results of 192 COVID-19 patients who had continuous serological detection during hospitalization was conducted. The results of some patients returning to hospital during the rehabilitation period were also included. We observed that the SARS-CoV-2 IgM seroconversion time of most patients was 5-10 days after the symptoms onset, and then rose rapidly, reaching a peak around 2 to 3 weeks, and the peak concentration was low (median concentration 2.705 AU/mL). The duration is short (within about one week), and then enters the descending channel. IgM has not been detected in one third of the patients about 5 weeks after the symptoms onset, and IgM has not been detected in more than half of the patients around 8 weeks, which indicated that serological outcomes have occurred. IgG seroconverted later than or synchronously with IgM, reaching peaks around 3 to 4 weeks.The peak concentration was higher than that of IgM (median concentration 33.998AU / ml).The peak duration last for a long time, about 1-2 weeks, and then entered into a plateau or slowly decreasing, the duration can be as long as several weeks. IgG against SARS-CoV-2 could be detected in 96% of patients at about 8 weeks after the symptoms onset, and the median concentration was high at 20.977AU/mlL. This is consistent with the antibody production rules we have known in the past: IgM seroconvert early, with low concentration and short maintenance time; IgG seroconvert late with high concentration and long maintenance time. The results from our observations can also be inferred: for the detection of double serum in the acute phase and the recovery phase of suspected patients with COVID-19, IgG titer increased by 4 times or more has a definite diagnosis.

As some founding in other literature [9, 10], IgG seroconvert earlier than IgM was found in 34% of COVID-19 patients in our study. We believe that if the statistical results alone infer that IgG seroconvert earlier than IgM, there may be a risk of misjudgment. We consider that the reason may be related to the characteristics of low concentration and short existence time of IgM. In addition, the patient had a long time to seek medical advice after the symptoms onset, therefore it was impossible to monitor the seroconversion in patients during this time. Due to the limitations of the study, it is difficult for us to observe the results of daily blood sampling after admission to the hospital, which will inevitably cause missed IgM seroconversion. This is also one of the limitations of the research.

The first serological blood samples were drawn from COVID-19 patients within two days of admission. We sorted out the detection results of 3 time points (the first test point, the last negative point and the first positive point).The results of 3 time points were compared with the results of the control group, we found that the concentration of antibodies in patients before the seroconversion was higher than that of healthy population, with an increasing trend. It indicated that the antibodies of COVID-19 patients began to produce early in the infection, however the concentration of them have not reached the threshold we set so couldn’t be detected. Therefore, COVID-19 suspects whose concentration of antibodies are below “sea level” cannot be judged uninfected by only one test. The serological test of COVID-19 suspects should be dynamically observed multiple times to find out whether the antibody has an upward trend.

In the cohort of our observation, IgM and IgG of 13 patients were not detected, while 12 patients were observed during hospitalization to rehabilitation period, and only one patient remained negative during hospitalization, unfortunately missing observation in rehabilitation period. All of them are non-severe patients, 3 patients with asymptomatic infection (CT no pneumonia manifestation during hospitalization, all RT-PCR results were negative) and the other 10 patients were mild or common patients. We speculate that because of the extremely load of virus, the short exist duration and the virus eliminated fast by host, so the antibodies in these patients had low concentration and disappeared in a short time,which caused them could not be detected.

In the study, we also hope to understand the relevant factors of antibody production through statistical analysis of the clinical data of COVID-19 patients. Consistent with the results of multiple reports, we found that most of the severe patients are elderly and have a longer hospitalization time. Their antibody concentration is higher than that of the mild patients during the entire observation period. We found that the RT-PCR results in severe patients have been positive for a long time, and the IgG level has been maintained at a high concentration. The virus infection causes disease whether depends on the pathogenicity of the virus and the immune function of the host. If the load of viruses that invade the host is small and can not reach the target cells, or the virulence is weak, but the host has a strong anti-virus immune function, so the host can quickly wipe out the virus through the immune response. On the contrary, if the virus survives and proliferates in the host for a long time, as we have observed that the immune system of the host will always work, protective antibodies would also maintain at a high concentration level. We also found that IgM levels in males are higher than that in females, which may be related to the different immune function between male and female, and the related mechanisms are unclear and need to be explore in the future.

In addition, we began to screen 216 medical examination population and found that IgM or IgG in 7 cases were weakly positive. The concentration of IgM and IgG results were 0.300-1.575AU/mL and 0.325-5.002AU/mL, respectively. After various investigations and tests, 4 of them were found to be hepatitis B patients, 2 of then were syphilis patients, and 1 patient was Autoimmune Patients. These tests were performed multiple times, and IgM or IgG were weakly positive. We believe that these cases are false positive results, and the false positive rate is 3.2%.The existence of such false positives is determined by the characteristics of immunological testing, that is, antibody testing will result in false positives due to the presence of some interfering substances in clinical specimens. Common interfering substances are divided into endogenous and exogenous. Endogenous interfering substances generally include rheumatoid factor, heterophilic antibody, complement, mouse anti-Ig antibody due to treatment with mouse antibody, etc. The generation of exogenous interfering substances may caused by specimen hemolysis, contamination of the specimen with bacteria and the long storage time of the specimen. Therefore, when analyzing antibody results, we must pay attention to excluding the influence of the above interference factors. If dynamic observation is used, it can also help us find and exclude these false positives, and they will not show the peak curve of antibody production.

The research object we observed came from Guangzhou, China. A total of 505 cases of COVID-19 were diagnosed in Guangzhou. It was characterized by no widespread epidemic. Most patients had mild symptoms, short duration. The pattern of antibodies expression in Guangzhou may differ from the pandemic area. In this study, it was observed that antibodies against SARS-CoV-2 of COVID-19 patients in Guangzhou produced early after disease onset: IgM had lower concentration and short duration while IgG was higher in concentration and maintained for a long time. Antibody concentration is positively correlated with the severity of the disease and the duration of virus exist in host. A cohort of 107 patients in this study was returned to the hospital for serological testing one month after discharge. The positive rate of IgM was 39.3% and the positive rate of IgG was 89.7%. How long can IgG against SARS-CoV-2 last in the human body? Can the antibodies in the serum of the rehabilitation patients be used as protective antibodies? Do COVID-19 patients have secondary infection during antibody positive period after recovery? This is an issue that everyone is currently concerned about, and it is also the focus of our follow-up observation in the future.

## Acknowledgements

We acknowledge all health-care workers involved in the diagnosis and treatment of patients in Guangzhou Eighth’s People Hospital.

